# Self-Rated and Social Determinants of Health Spending Attitudes in the United States

**DOI:** 10.1101/2020.05.07.20094342

**Authors:** Mazbahul G. Ahamad, Fahian Tanin

## Abstract

Self-rated and social determinants of health are associated with people’s health attitudes towards government spending on improving national health in the US. We used data from eight biennial nationally representative General Social Surveys from 2004 to 2018 (n = 21116) to assess the determinants of health spending attitudes among US adults through a multivariate logistic regression technique. We found that more than three quarters of adults responded that government health spending was ‘too little’ for improving and protecting American health. We also found that adults those with ‘fair’ self-rated health were more likely to respond ‘too little’ or negative about health spending after adjusting for socioeconomic and sociodemographic factors and political regimes. This result suggests exploring individual-specific reasons associated with this negative attitude to restructure health spending priorities in the US.

## 1. Background

In the US, nearly three quarters of surveyed adults responded that the government was spending too little or negative on improving and protecting the nation’s health, during different political regimes in 2004—2018. This negative attitude varied between four self-rated health groups (e.g., excellent, good, fair, and poor) (1), which might explain their healthcare expectations and priorities (2). Health spending attitude often involves socioeconomic status and demographic characteristics (2–4). It is, therefore, reasonable to assume that attitude towards government health spending is contingent on individual health perceptions and associated determinants (5).

The association between health spending—related attitudes and self-rated health along with other social determinants (2) and current political regime (6), is complex and largely unexplored. To design priority-based public health programs, understanding the association between these determining factors is essential. In this paper, we investigated the determinants of heterogeneous health attitudes among the US adult population.

## 2. Materials and Methods

### 2.1. Data

We used data from eight biennial nationally representative General Social Survey (GSS) (1) from 2004—2018 in the US. We used individual-level but de-identified data people’s health spending attitudes and self-reported health variables, along with other sociodemographic, socioeconomic, and political regime-related indicators for statistical analyses.

### 2.2. Model variables

The primary outcome variable was a dummy variable representing an adult respondent’s attitude towards government health spending (1 if they responded ‘too little’ or negative, 0 otherwise). The main variable of interest was self-rated health condition, which was also a category variable (excellent, good, fair, or poor). We also included socioeconomic and demographic variables such as age (18–44, 45–64, >64 years), gender (male or female), education (high school, college, or >college), place of growing up (farm or non-farm), income (<$10,000, $10,000—$25,000, or >$25,000), working status (working or otherwise), and political regime (2004—08 and 2018 as regime #1, and 2010—2016 as regime #2).

### 2.3. Statistical analysis

We used logistic regression models to estimate odds ratios (ORs) with 95% confidence intervals (CIs) to investigate the associations between the primary outcome (health spending attitude) and main variable of interest (self-rated health), controlling for different sociopolitical characteristics. The level of statistical significance was specified as p ≤ 0.05. We used Stata version 16.1 for both descriptive and statistical analyses (7).

## 3. Results

During 2004–18, most adults (68.59%) rated government spending on improving and protecting the nation’s health as too little. The smallest proportion of respondents rated their health as poor (5.13%), 20.05% as fair, 47.60% as good, and 27.23% as excellent. Across all self-rated health groups, the ‘too little’ or negative health spending attitude was high, ranging from 68% to 70%. Both variables showed fluctuating trends during the same period. Approximately 36% of respondents were aged 45-64 years, around 54% were female, and 33% grew up on a farm until 16 years old (Table 1). Half of the participants had at least college-level education, only 58% earned more than $25,000, and 66% reported they were working. Table 1 presents multivariate logistic regression results with the distribution of model variables. Of 21116, 2881 responses (13.64%) were included for statistical analysis. We found that adults who reported fair health were 92% more likely to rate government spending on health as ‘too little’ or negative compared to the excellent health group (OR = 1.92, p = 0.00), after controlling for age, sex, place of growing up, education, income, working status, and political regime.

**Table 1.** Distribution and Associations Between Health Attitudes and Self-Rated Health Along with Social Determinants in the US, 2004–2018.

| Variable | Health attitudes* |  |
| --- | --- | --- |
|  | Distribution** | OR (95% CI) |
| **Self-reported health** |  |  |
| Excellent | 3626 (27.23) | 1.00 |
| Good | 6623 (47.60) | 1.38 (0.81, 1.96) |
| Fair | 2912 (20.05) | 1.92 (1.24, 2.99) |
| Poor | 809 (5.79) | 2.03 (0.49, 8.37) |
| **Age, year** |  |  |
| 18-44 | 9642 (48.98) | 1.00 |
| 45-64 | 7380 (35.55) | 0.78 (0.55, 1.12) |
| >64 | 3854 (15.47) | 0.51 (0.30, 0.86) |
| **Gender** |  |  |
| Male | 9458 (45.88) | 1.00 |
| Female | 11658 (54.12) | 1.67 (1.00, 2.08) |
| **Grown-up place** |  |  |
| Farm | 9148 (45.58) | 1.00 |
| Small city | 6491 (32.51) | 1.07 (0.72, 1.57) |
| City | 3941 (18.91) | 0.80 (0.51, 1.28) |
| **Education** |  |  |
| High school | 8980 (42.09) | 1.00 |
| College | 9133 (44.37) | 0.97 (0.67, 1.40) |
| >College | 2911 (13.54) | 0.98 (0.59, 1.64) |
| **Income, \$** | | |
| <10,000 | 2969 (24.62) | 1.00 |
| 10,000-25,000 | 2031 (16.60) | 0.97 (0.54, 1.73) |
| >25,000 | 7312 (58.78) | 0.75 (0.50, 1.14) |
| **Working status** |  |  |
| Working | 7763 (34.08) | 1.00 |
| Otherwise | 13335 (65.92) | 0.71 (0.41, 1.22) |
| **Political regime** |  |  |
| Regime #1 | 11771 (55.74) | 1.00 |
| Regime #2 | 9345 (44.26) | 0.34 (0.24, 0.49) |

*Notes:* \*The primary outcome (dependent) variable was a dummy variable representing an adult respondent's attitude towards government health spending (1 if they responded 'too little' or negative, 0 otherwise). \*\*The distribution presents unweighted numbers and weighted percentages.

## 4. Discussion

Our findings show that the perception of too little government spending to improve and protect the nation’s health was common among all four self-reported health groups. This finding is consistent with the results of a previous study (8). From the regression analysis, however, we found that those with fair self-rated health were more likely to rate spending as too little, after adjusting for socioeconomic, sociopolitical, and political regime characteristics. We also found similar results without adjusting for any sociopolitical characteristics. We found no statistically significant relationship between excellent, good, or poor self-rated health and attitude of too little government health spending. However, these results do not necessarily imply an absence of possible relationships between all four self-reported health statuses and their health spending attitudes. These results suggest that health spending attitude might depend on other sociopolitical concerns such as attitude to spending on foreign aid (9). As related research has suggested, political ideology and trust likely contribute to shaping these attitudes (10). Future studies should evaluate whether health spending attitude intersect with other variables such as race, ethnicity, and political involvement to rigorously explain the effect of self-reported health on health spending attitudes.

One of the major limitations of this study was self-reported health and perception data, which may be subject to reporting bias. We need to be cautious in generalizing the results with any causal mechanisms and the results may be limited to the sociopolitical controls related to the study. Despite the limitation, our descriptive results shed light on the high prevalence of negative health spending attitude among US adults in a social context. The study contributes to the empirical literature on self-rated health, sociopolitical determinants of health, and health spending attitudes of US adults in improving and protecting the nation’s health.

## 5. Public Health Implications

Self-reported fair health was associated with a perception of too little government health spending. Public health policies and program interventions should focus on additional understanding of the social and political causes of people’s negative perceptions to improve structural disparities in national health spending priorities. As the government can be a part of health outcome improvement programs (11), priority-based resource allocation on health spending is important.

## Data Availability

Ethical approval was not sought as we used publicly available data from GSS (https://gssdataexplorer.norc.org).

## Authors’ contributions

MCA planned the study and extracted data from GSS. MGA performed statistical analysis, data interpretation and wrote the first draft of the manuscript.; then MGA and FT contributed to writing and editing of the manuscript. All authors have access to the data, codes, and revised all drafts to produce the final version of the paper.

## Acknowledgments

We appreciate GSS at the University of Chicago for providing data for the study.

